# Prediction of 2019-nCov in Italy based on PSO and inversion analysis

**DOI:** 10.1101/2020.05.08.20095869

**Authors:** Sun Siyi, Zheng Yangping

## Abstract

Novel coronavirus (2019-nCov) has swept the world, and all of the world have been harmful. This article makes prediction and suggestions for the Italy. Up to March 11, 2020, 2019-nCov thoroughly broke out in Italy with over 10,000 confirmed cases notwithstanding the gradually block of the country since March 9, 2020. Estimation of possible infection population and prospective suggestion of handling spread based on exist data are of crucial importance. Considering of the biology parameters obtained based on Chinese clinical data in Wuhan, other scholars’ work and real spread feature of 2019-nCov in Italy, we built a more applicable model called SEIJR with log-normal distributed time delay to forecast the trend of spreading. Adopting Particle Swarm Optimization (PSO), we estimated the early period average spreading velocity (***α***_**0**_) and conducted inversion analysis of time point (***T***_**0**_) when the virus first hit the Italy. Based on fixed ***α***_**0**_ and ***T***_**0**_, we then obtained the average spreading velocity ***α***_**1**_ after the lock by PSO. For the aim of offering expeditious advice, we generated the prediction trends with different ***α*** which we considered would be helpful in addressing the infection. Not only solved the complex, nondifferentiable equation of epidemic model, our research also performs well in inversion analysis based on PSO which conveys informative outcomes for further discussion on precatious action. To conclude, the first day of spread is around February 1, 2020 with the early period average spreading velocity *α*_0_=0.330 which is higher than most cities in China except Wuhan. After locking the country and attaching great attention to public precaution, the *α*_1_ sharply descended to 0.278, indicting the effectiveness of these measures. Furthermore, in order to cope the disease before mid-April, take actions to control the under 0.25 is necessary. Code can be freely downloaded from https://github.com/Summerwork/2019-nCov-Prediction.

## 1 Introduction

A global epidemic disease known as the novel coronavirus (2019-nCov) had seriously hit the most area around the whole world causing unpredictable loss of manpower and finance during the first quarter of 2020 [21] [1]. The first confirmed 2019-nCov case was reported in Wuhan, China on December 31, 2019 (World Health Organization, 2020a). As of March 19 (21:00 GMT),2020, 2019-nCov has resulted in 81262 confirmed cases and 3250 dead cased in China cumulatively (National Health Commission of the People’s Republic of China, 2020). China as the first country faced by the outbreak of the severe disease, it took strict but effective action to contain the spread of 2019-nCov and attained apparent success till now. [21] Many related works have been done in prediction and precaution via constructing proper model and analyzing parameters. [21] [20] [19]

While in the other parts worldwide, the menacing disease just became to spread [7], especially in countries with no preparation and experienced measures for suppressing the possible large-scale infection. In this article, we take Italy which is now experiencing severe situation of 2019-nCov as example to conduct analysis with the aim of offering utilizable suggestion. In the first period (before March 9, 2020), we attempt to inverse the virus spread timeline in Italy and the early period average spreading velocity by adopting PSO to optimize the parameters based on our SEIJR model and existing data (European Centre for Disease Prevention and Control). In the second period (only use data from March 9-16, 2020), we optimize the average spreading velocity in order to show the effect of country blockade. In the last period of our research, we demonstrate latent trends of confirmed cases with various average spreading velocity which of vital importance in controlling the disease. The whole flow chart is illustrated in Figure 1.

**Figure 1.**
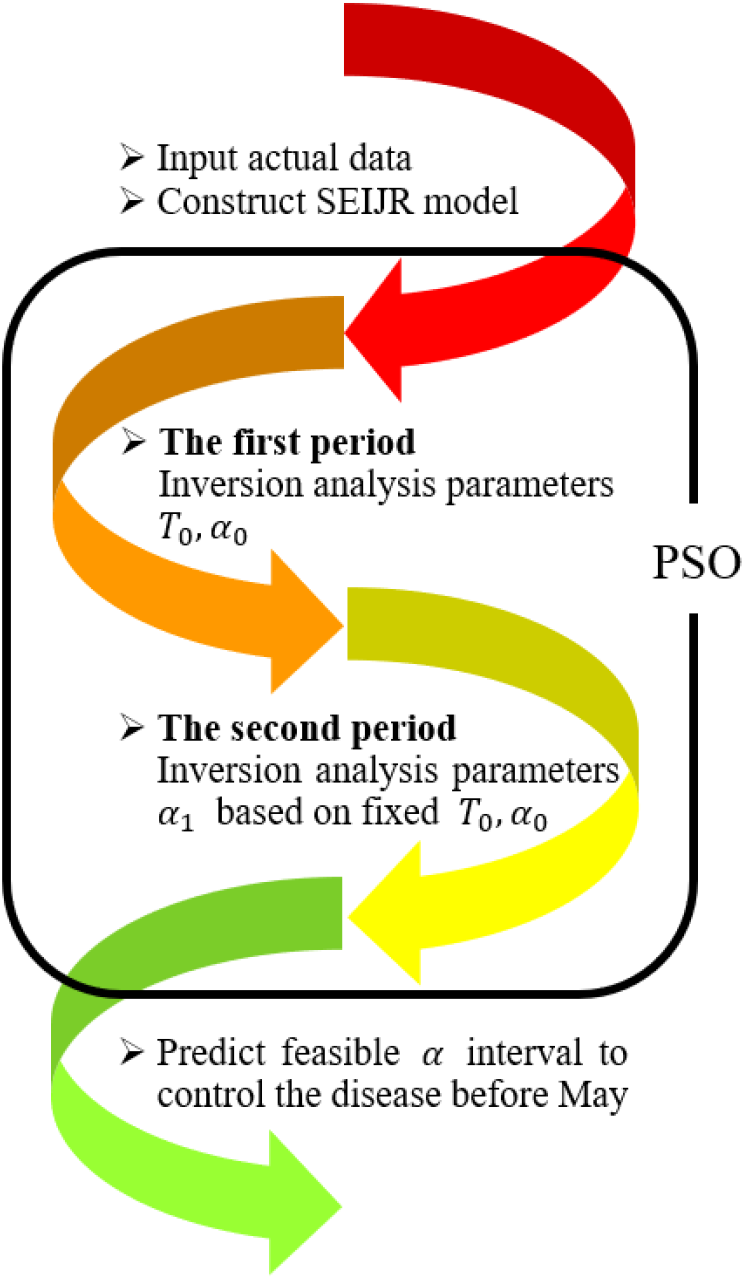
Flow chart

## 2 Data and Model

### 2.1 Data Collection and Processing

We collected the daily reported confirmed diagnosed data from the website of European Centre for Disease Prevention and Control (European CDC). All these data is public for everyone.

Based on the need of our analysis, we preprocessed the data by adding the daily reported confirmed diagnosed cases to obtain the accumulative amount for following inversion of parameters and prediction. Here are details about the processed data we used in this article (Table 1).

**Table 1.**
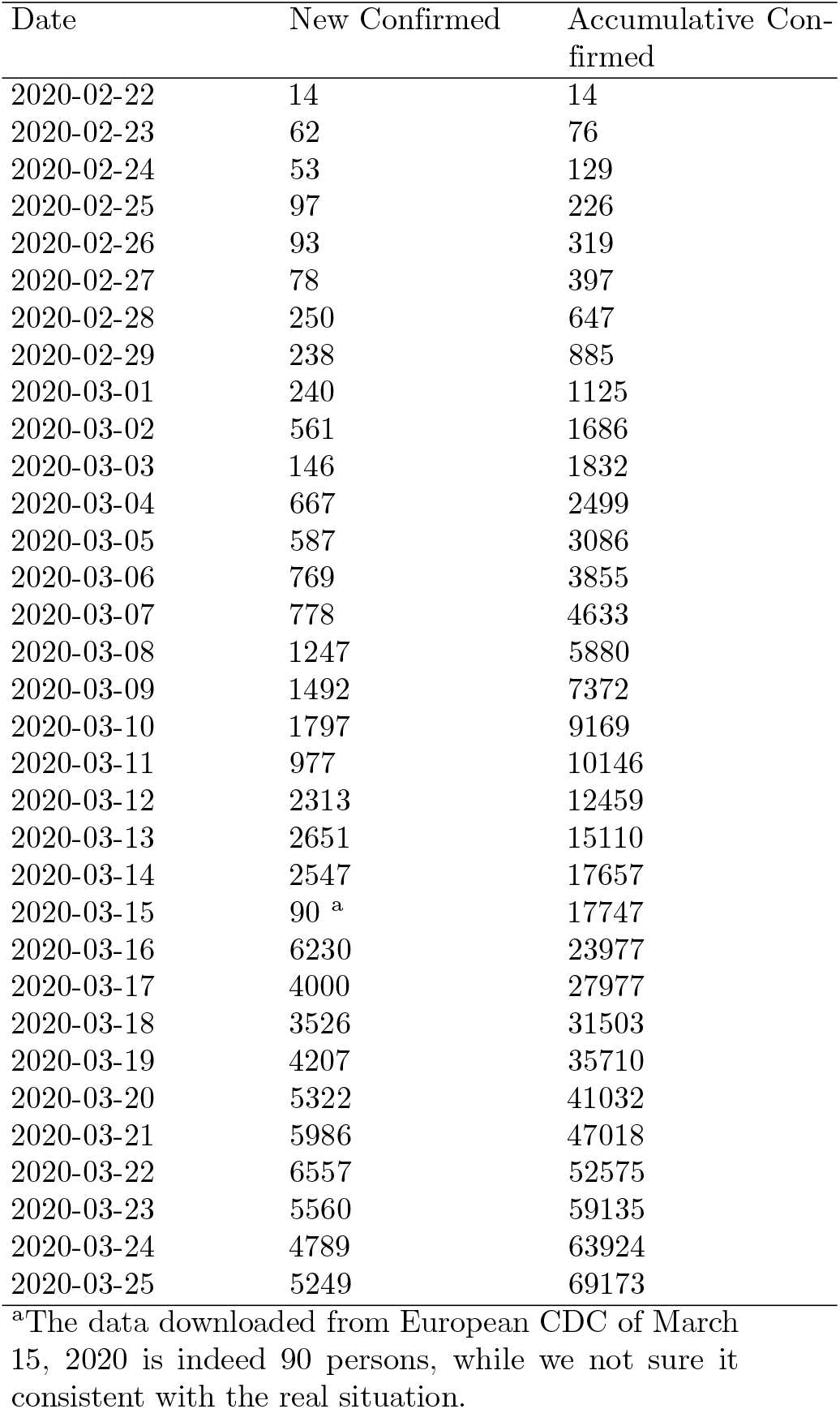
New Confirmed and Accumulative Confirmed Data

### 2.2 SEIJR Model

Disease transmission is a complex process with multiple variables and uncertainties making it unable to be accurately solved and predicted [16], [11] [6]. However, models are feasible in forecasting for infectious diseases when different characteristics parameters [18] [17] like transmission mode, immunization mode, mortality and average spreading velocity are offered. Classical models for infectious diseases include SIR model [2] and SEIR model, etc. Considering the actual situation in Italy and the transmission characteristics of 2019-nCov obtained in China, this paper built the SEIJR model with log-normal distributed time-delay terms [15] [3]based on the SEIR model [14] [12]. Figure 2 illustrates the SEIJR model. The model describes the problem by assuming population consists of six types (accumulative value): susceptible population *S*, exposed population *E*, infectious population *I*, confirmed *J*, recovered population *R* and dead population *D. α* is average spreading velocity, *β* is diagnose rate, *γ*_1_, *γ*2 are die rate and is cure rate. *t*_1_ (*t*) is the time of incubation period [5] *E* need to become *I, t*_2_ (*t*) is the time of waiting period *I* need to become *J* and *t*_3_ (*t*) is the duration of hospitalization [5] *J* need to become *R. S*(*t*), *E* (*t*), *I* (*t*), *J* (*t*), *R*(*t*) and *D* (*t*) are dependent variables of time *t*, respectively. *α, β, γ,μ*are constants only related to the actual situation. Time-delay *t*_1_,*t*_2_,*t*_3_ are only depend on time *t*, respectively, which can be written as *t*_1_ (*t*), *t*_2_ (*t*) and *t*_3_ (*t*).

**Figure 2.**
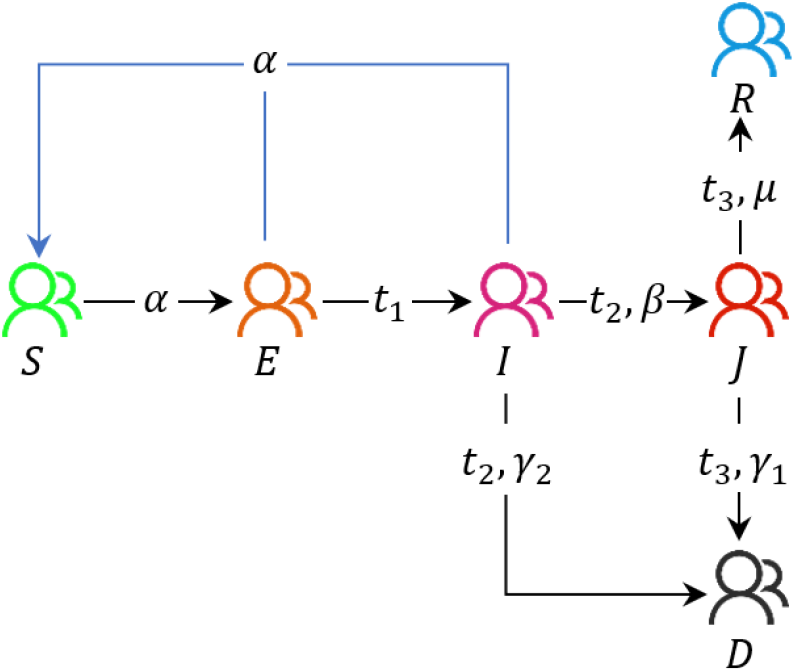
SEIJR model

### 2.3 Interpretation

In the model, only *E* and *I* have ability to infect *S* which is a process of contact infection with transient time. *E* indeed infected but shows no symptoms of 2019-nCov and then transfers to *I* after an incubation period *t*_1_. *I* has symptoms like fever, cough and shortness of breath. Because the pre-virus symptoms are not obvious [5] and the uneven medical facilities in Italy, *I* will be confirmed as *J* after a period *t*_2_. Due to the seriousness and infectivity of the virus, it can be considered that when it becomes *J, J* will be immediately isolation and lose its ability of infection. Treatment will be started immediately after confirmed. *J* will recovery and become *R* after a duration of hospitalization *t*_3_. Because of the 2019-nCov has certain lethality, it needs some more assumptions:

#### 2.3.1 Assumption 1

The number of deaths during the period of *E* is too tiny to be considered, only need to consider the mortality during the period of *I* and *J*;

#### 2.3.2 Assumption 2

The data during the period of *I* is unavailable. We consider that *I* to *D* and *I* to *J* have the same delay time *t*_2_ which means the only difference between them is proportion;

#### 2.3.3 Assumption 3

The official organization and medical institutions do not give any information on how long for patients in the treatment stage will die, but there exists clinical information that for a confirmed patient how long the patient is needed to be cured [5]. As the same way *J* to *R, J* to *D* have the same delay time *t*_3_ only with different proportion.

People in *I* can be diagnosed and receive treatment with *β* diagnose rate. Otherwise, they will die with the die rate *γ*_1_ = 1−. The recover rate for *J* is and the dead rate is *γ*_2_ = 1 −*μ*. The total mortality proportion is *γ* = *γ*_1_ + *γ*_2_.

#### 2.3.4 Assumption 4

After recovering, people in R will not go out because they are in a frail state and will be considered as isolation.

### 2.4 Time-delay Function

Combining the basic principles of epidemiology and etiology, it can get that the time *t*_2_ from infection to diagnosis, which approximately follows the log-normal distribution [8]. The assumption can be applied to *t*_1_ and *t*_2_ without loss of generality. For *t*_1_, due to the lack of Italian clinical information statistics, it’s more accurate and reliable to use the clinical data of Zhong NS et.al: the median incubation period is 4 days [5], the quartile is 2 and 7 days. With the log-normal distribution’s formula:

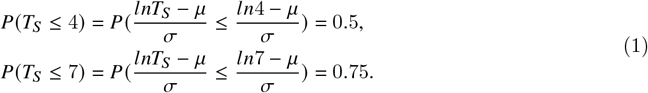

Get 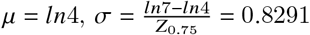 where *Z*_0.75_ is normal distribution quartile. Then, the log-normal density function corresponding to *t*_1_ is:

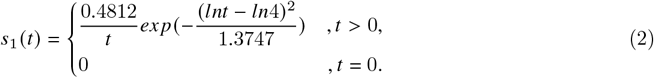

For *t*_2_, the data in Italy is still unavailable, but the relevant distribution function is given in the Chinese research report. In the paper of Yang ZW et.al, the relevant statistics of “the time interval between the most recent stay in Hubei Province and the confirmed diagnosis” are given [4]. The end point of this period corresponds to the start point of *J* in the SEIJR model, but there is no corresponding start point. It can only be known that it’s in the middle of the time between *t*_1_ and *t*_2_, it is convincing to regard it as the point when *e* enters *I*, which means the time given in that article corresponds to the *t*_2_ in this article. As the same way, the log-normal density function corresponding to *t*_2_ is:

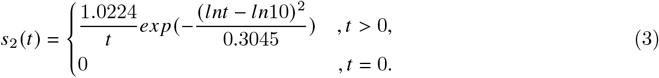

For *t*_3_, the Italian official organizations and medical institutions still lack clinical information and official statistics, so we also assumes that *t*_3_ follows the log-normal distribution. However, Zhong NS et.al gave some relevant data: the duration of hospitalization which is *t*_3_, the median is 12 days, and the quartiles are 10 and 14 days [5]. Adopting the same method used in *t*_1_ estimation, we can get the distribution density function corresponding to *t*_3_ is:

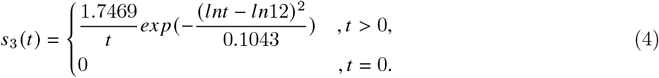

Now give the definition of log-distributed normal time-delay function in SEIJR model:

#### Definition 1 (Time-delay Function in SEIJR Model)

The time-delay function describes how many people change this stage from the previous stage at time t

The general formula is written as:

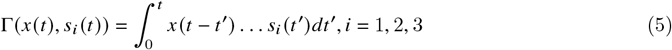

where *x* (*t*) is the term which has time-delay, and *s*_*i*_ (*t*) is the distribution density mentioned before. Here we finally get the precise differential equation of SEIJR model as:

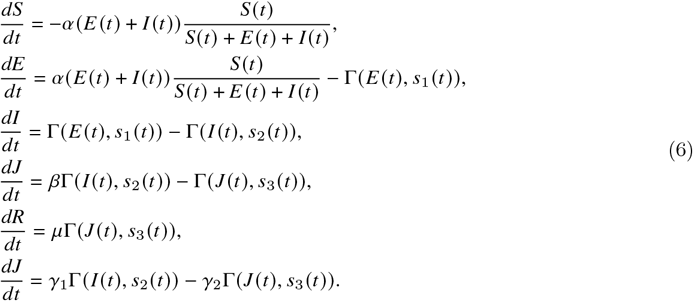

## 3 Methods

### 3.1 Runge-Kutta Methods

The previous differential Eq. (6) corresponding to the SEIJR model is a form with integrals and independent variables on the integral limit which from the time-delay function term,so there is no analytical solution. For this case, numerical methods are useful. We combine iteration and degree four Runge-Kutta to generate the numerical solution with the initial condition. Here is the main principles of Runge–Kutta fourth-order method [8], let the differential equation have the form as follow:

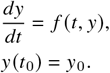

Then its iterative formula is:

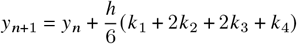

where

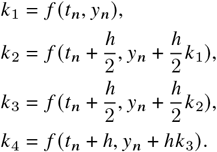

### 3.2 Particle Swarm Optimization

Usually the epidemic model’s descriptive function was derivative-based without considering time delay or just assume fixed linear time delay, such as SIR model. In this article, the SEIJR model solved by combining iteration and degree four Runge-Kutta mentioned before. Accounting for this, the function of least square method (LSE) employed in addressing this problem for different periods are denoted as follow:

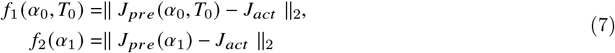

where *f*_1_ is LSE of the first period. *f*_2_ is LSE of the second period. 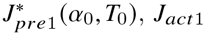 are predicted and actual value from February 22 to March 9, 2020. 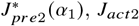 are predicted and actual value from March 9 to March 25, 2020. Both of them are nonlinear, complex, discontinuous and nondifferentiable [13]. Traditional method based on derivation is infeasible for minimization. While approaches such as annealing algorithm [10] to search for parameters is more calculative expensive than the particle swarm optimization (PSO).

PSO [14] is a population-based search algorithm sparked by the forage behavior of birds within a flock. Individuals gain the ability of searching for better solution areas by learning the fitness information from the environment. In PSO algorithm, the velocity of individual is dynamically changed considering its previous flying experience.

The algorithm consists of three main parts: individual best, global best and individual optimization based on the best particle of whole population [9]. In this article, *f*_1_ and *f*_2_ are the fitness function during the first and second period. The main process of conducting this algorithm and basic parameters are respectively shown in Figure 3 and Table 2.

**Table 2.** Parameters for PSO

| Parameter | Value |
| --- | --- |
| $c_1$ | 2 |
| $c_2$ | 2 |
| $\omega$ | 0.6 |

**Figure 3.**
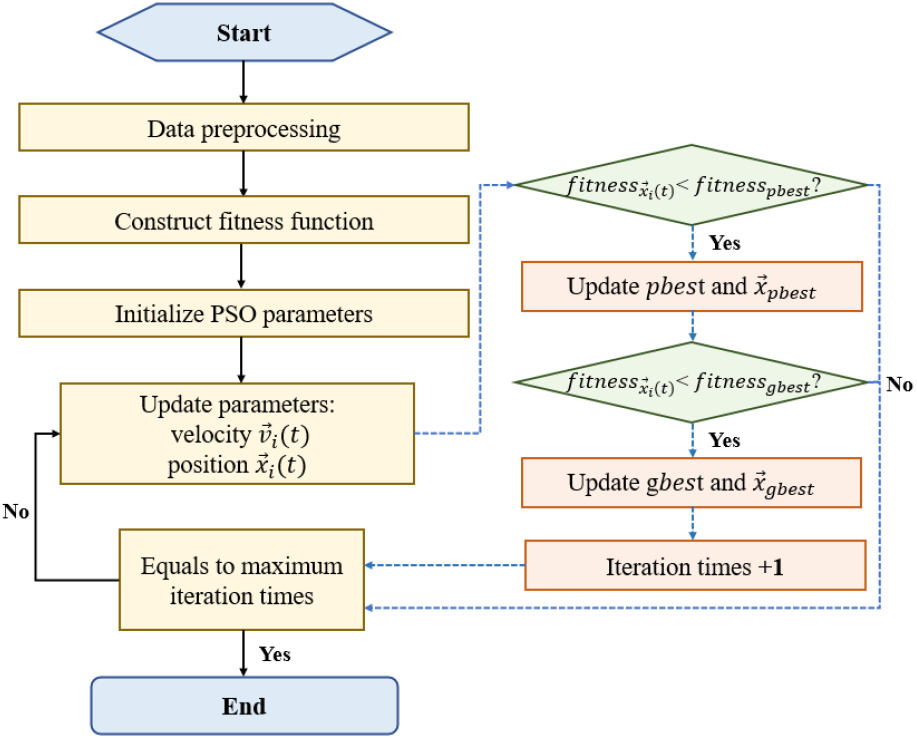
PSO flow chart

## 4 Results

### 4.1 The first period

During this period of our work, we adopt PSO using *f*_1_ as its fitness function to generate the optimal *α*_0_,*T*_0_. As shown in the Figure 4, the best result is *T*_0_=21 (after rounding) and *α*_0_=0.33. After obtained the greatest *α*_0_,*T*_0_, we draw the prediction curves for further comparison in the following section. Illustrated in Figure 5a is the prediction value 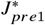 with the optimal parameter and the actually value *J*_*act*1_; (b) is the same curve with whole actual value *J*_*act*_ if the average spreading velocity remain unchanged.

**Figure 4.**
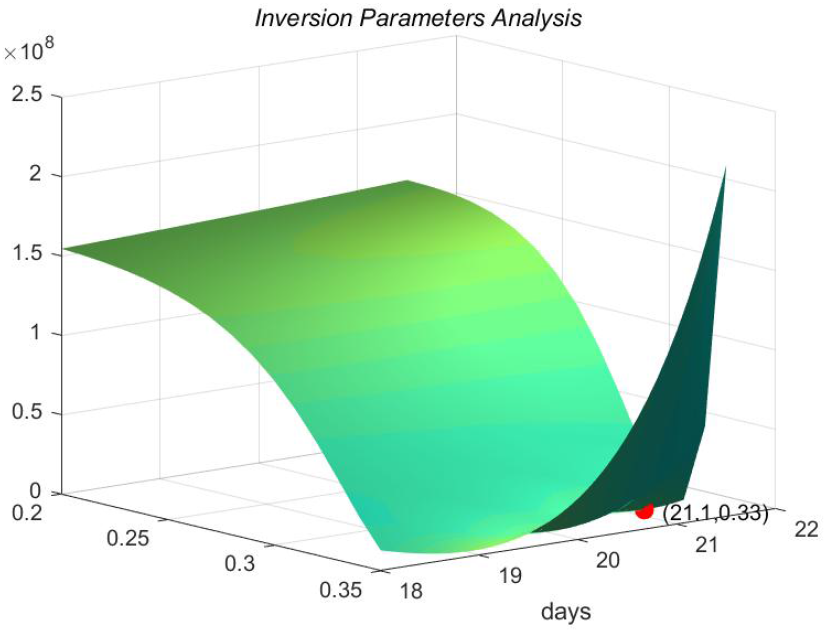
Vistualization of PSO searched results

**Figure 5.**
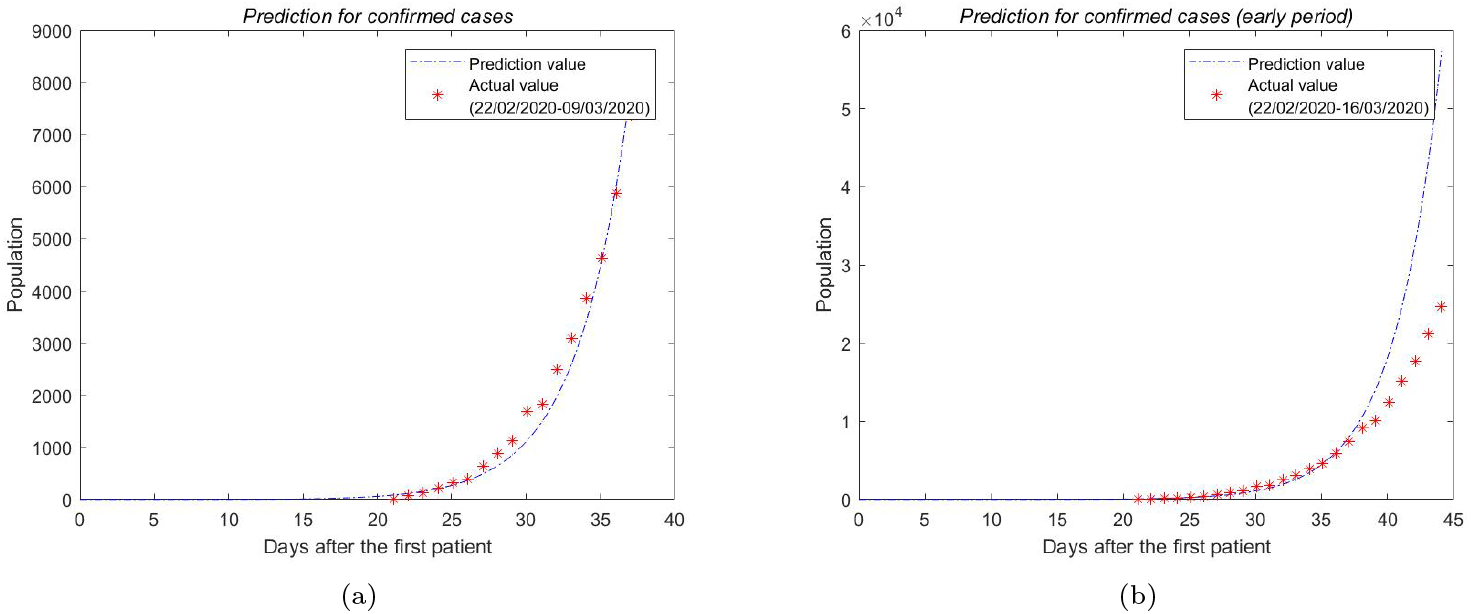
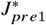 and *J*_*act*1_,*J*_*act*_ with *α*_0_=0.330

### 4.2 The second period

Considering of the precautious action like blockade taken by Italy government and the information Figure 5b conveyed, we assumed the average spreading velocity changed after these actions. Based on fixed *α*_0_,*T*_0_, we applied PSO to the optimization problem and got the result of *α*_1_=0.278 which is cut down from *α*_0_. The Figure 6 shows the predicted population 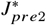 with *α*_1_ from March 9 to 25, 2020 and *J*_*act*2_.

**Figure 6.**
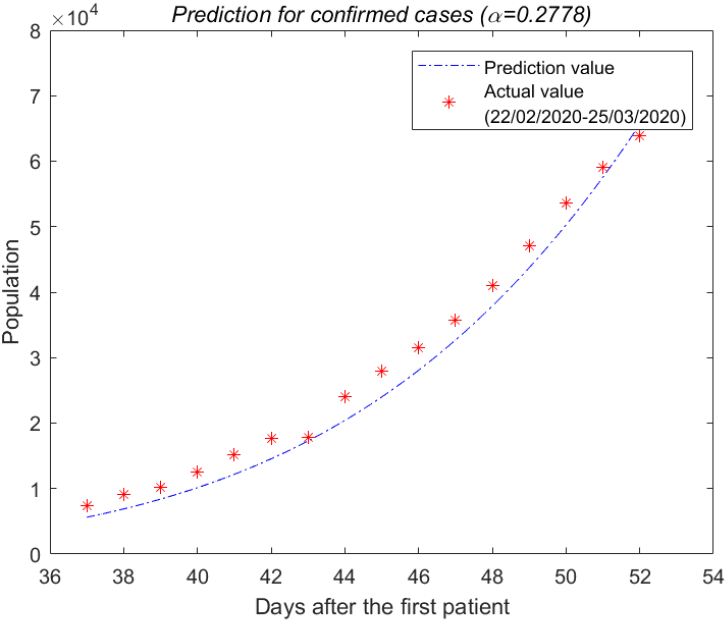
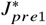 and *J*_*act*2_ with *α*_0_=0.278

### 4.3 Predicted J trends with different *α*

Figure 7 gives prediction curves with different *α* with the aim for further discussion about controlling the disease before mid-April.

**Figure 7.**
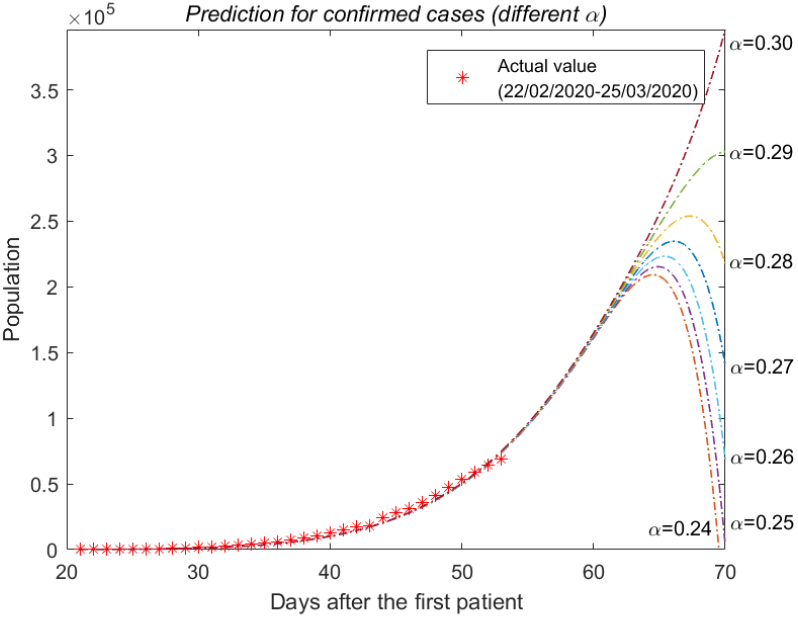
*J*_*pre*_ with different *α*_0_

## 5 Discussion

### 5.1 Reliability of SEIJR model

It is clearly seen that the fitting curve of *J* in the SEIJR model fits well with the confirmed diagnosis data of the Italian official statistics. *T*_0_ = 21 represents the initial value of the model which indicates the first *E* appeared 21 days before February 22 that is February 1. According to the information reported by the Italian government, the first case in Italy, when *J* appeared, was January 31. Two patients from Wuhan, China arrived in Italy by air at January 23 and visited other cities in Italy. They finally arrived in Rome, feeling physical discomfort at January 30 and was confirmed diagnosis then isolated at January 31. It is reasonable to speculate that they had carried the 2019-nCov in Wuhan before arriving in Italy in January 23, and continued to spread in Italy for eight days after January 23 until January 31. The first case of *E* reported in official data appeared at January 23, and the model’s conclusion appeared at February 1. In fact, because the two senior travelers are 67 and 66 years old with poor mobility, unfamiliar surroundings and the simple factors of interpersonal relationships, they have a lower *α* which also leads to real initial time point is earlier than the initial time point of the theoretical model. To sum up, the model has higher accuracy and stronger credibility. [11]

### 5.2 Effect of blockade

Prime Minister Giuseppe Conte extended the quarantine lockdown to cover all the region of Lombardy and 14 other northern provinces on March 8, and all region on March 10. At the same time, the Italian government further banned rallies and sports activities nationwide, announced a national blockade, and unnecessarily stopped going out. Compared the average spread velocity *α*_1_ = 0.278 with *α*_0_ = 0.330 at the early stage(before March 9, 2020), a decrease existed after the series of actions which verifies the effectiveness of this measure.

### 5.3 Advice to Italy government

According to the model’s prediction, when *α* is in a suitable range, the number of confirmed patients will gradually decrease. It is obvious that the smaller the value of *α* is, the faster the number of confirmed diagnoses decreases. Since Italian government hopes to end the 2019-nCov by mid-April, it can control *α* around 0.25 according to the prediction of the model. Based on this conclusion, Italy government is supposed to strengthen the isolation, reduce gathering activities and make people understand the importance of precautionary measures to constrain *α* under 0.25.

## 6 Conclusion

Applying PSO to our SEIJR model with log-normal distributed time delay, we obtained the convincing start time (around February 1, 2020) of 2019-nCov and the average spreading velocity (*α*_0_=0.330) at the early stage. We compared the average spreading velocity during the early period and following period, a conspicuous decrease attributed to the effective measures was found. Based on the prediction interval of possible infected population of different *α*, we strongly recommend Italy to keep *α* under 0.25 if they want the situation take a turn for the better even ended before mid-April.

## Data Availability

We collected the daily reported confirmed diagnosed data from the website of European Centre for 51 Disease Prevention and Control. All these data is public for everyone. 52 Based on the need of our analysis, we preprocessed the data by adding the daily reported confirmed 53 diagnosed cases to obtain the accumulative amount for following inversion of parameters and prediction. 54 Here are details about the processed data we used in this article.

https://www.ecdc.europa.eu/en

## Acknowledgment

The authors would like to thank Dongmei Ai and Yan Xu at University of Science and Technology Beijing for helpful suggestions and discussion.

